# The assembly effect: the connectedness between populations is a double-edged sword for public health interventions

**DOI:** 10.1101/2020.05.18.20106161

**Authors:** Sai Thein Than Tun, Daniel M. Parker, Ricardo Aguas, Lisa J. White

## Abstract

Many public health interventions lead to disruption or decrease of transmission, providing a beneficial effect for people in the population regardless of whether or not they individually participate in the intervention. This protective benefit has been referred to as a herd or community effect and is dependent on sufficient population participation. In practice, public health interventions are implemented at different spatial scales (i.e. at the village, district, or provincial level). Populations, however defined, are frequently connected to other populations and this connectedness can influence potential herd effects. In this research we model the impact of a public health intervention (mass drug administration for malaria), given different levels of connectedness between similar populations and between populations of varying epidemiology (i.e. baseline transmission levels and intervention coverage). We show that the way such intervention units are connected to each other may influence the impact of the focal interventions deployed in both positive (adding value to the intervention) and negative (reducing the impact of the intervention) ways. We term this phenomenon the “assembly effect” which is a meta-population version of the more commonly understood “herd effect”. We conclude that public health interventions should consider the connectedness of intervention units or populations in order to achieve success.

## Background

The communicable diseases made up 44% and 31% of mortality in low and low-middle income countries.^1^ Public health interventions have been used for prevention of communicable diseases. Whenever these interventions reduce transmission of a communicable disease, there can be a community-level effect (“herd effect”).^2^ This herd effect provides a protective benefit to all members of a population, regardless of individual participation in the intervention.

Herd effects have been documented for any interventions that reduce the transmission potential such as early detection and treatment of pulmonary tuberculosis, mass drug administration (MDA) against lymphatic filariasis^2^, insecticide treated nets (ITN) against malaria infections^3^ and recently for MDA against *Plasmodium falciparum* malaria.^4^ Herd effects depend on sufficient population adherence to an intervention in order to provide a protective benefit to all individuals in the population. This threshold of participation has been considered in the context of a single population, with little consideration of the existence of meta-populations. Here, we explore how connectedness with other populations from different areas influences the effectiveness of the public health interventions, by giving malaria elimination as a working example.

Experts agree that global malaria elimination is achievable.^5-7^ Progression from malaria control to malaria elimination is a continuous process with different countries, subnational areas and communities at different stages on the pathway towards malaria elimination.^8^ To address the uneven landscape of malaria transmission in different areas, risk maps can be estimated, followed by a stratification algorithm to allow for better targeting and improved efficiency of malaria interventions.^8-10^ Despite having larger impact by only intervening in high risk areas, we must stress that these discrete areas may be connected to other areas of different malaria intensities, through human and/or mosquito movement. Population movement survey done in the Thai-Myanmar boarder area found that 44% of participants in one malaria cluster crossed the international boarder at least once a month.^11^ The two countries have different healthcare infrastructures and malaria transmission intensities^11,12^ and such connectedness could negatively impact the malaria elimination efforts on one side provided that no similar effort is made across the border. Previous models have also suggested the importance of human movement for efficient deployment of malaria interventions.^13,14^

We present a theoretical framework with two interconnected populations, hereafter referred to as “patches”. We explore how the magnitude of their connectedness impacts the potential success of MDA deployment in each of the patches. We related this to a documented example^4^ and predicted the outcomes of a series of alternative scenarios of connectedness, relative coverage, and relative transmission levels to obtain a more complete picture of this phenomenon and its implications.

## Methods

All simulations and analyses were carried out using the R software version 3.6.0^15^ with the following packages: deSolve,^16^ Rcpp,^17^ and lattice.^18^ A two-patch model was developed as an extension of a previously published single-patch model.^19^ Each patch is represented by an extended Susceptible-Infectious-Recovered-Susceptible (SIRS) compartmental model to incorporate the temporary effect of antimalarial drug action following MDA. The two patches are represented graphically as two intersecting circles (Figure 1), and force of infection (l) is defined in equation 1:

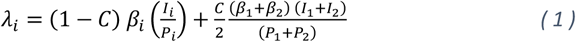

where *P* is the total population.

**Figure 1:**
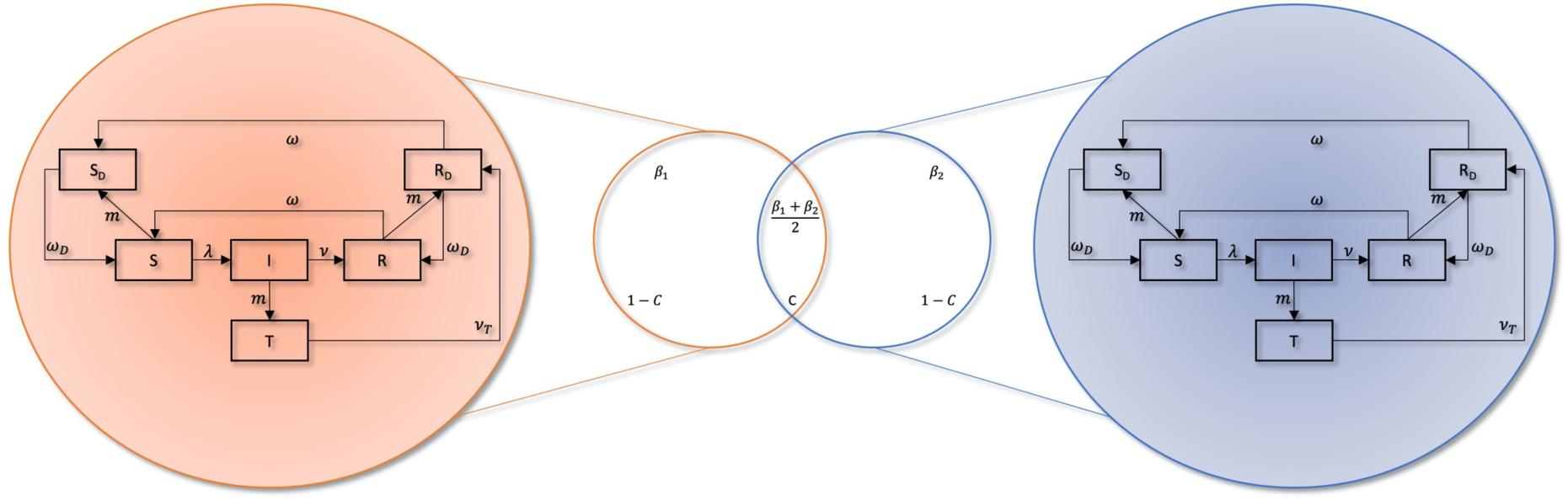
Two-patch compartmental model. C indicates the proportion of the population in each patch that shares a common infectious reservoir with the other patch. When the two patches are isolated (i.e. not at all connected, C=0), they share no infections and each individual’s risk of infection in a patch is completely independent of that in the other patch. At the other extreme of the connectedness spectrum (C=1), all individuals in the two patches are subject to the same force of infection (λ). β is the effective biting rate adjusted by vector interventions. Zoomed in areas describe the simplified compartments within each patch- S: Susceptables. I: Infected and Infectious; subgroups of I to capture different detactability and infectiousness are explained in the supplementary material. R: Recovered. T: Treatment. Compartments with subscript D denotes the temporary protection by having drugs.

MDA was modeled with a time-dependant rate *m* so that when MDA was implemented, a certain proportion of the population under coverage receive protection from the disease for some duration. The details of the model structure and interventions are in the supplementary material.

### Simulations

The two-patch model is simulated for several scenarios where one parameter of interest is varied at a time. The outcome metric measured from each patch in each simulation was whether a malaria elimination threshold, defined as “less than 1 infection per 1000 population per year”,^8^ was achieved one-year after the completion of a three-month MDA campaign. Since there are two patches, there are four possible outcomes: achieving malaria elimination in none of the patches, in patch 1 only, in patch 2 only and in both patches.

The results are plotted on a two-dimensional surface plot, where each axis represents the exploratory variable. On the X-axis, the connectedness parameter is increased from 0% to 100% with 1% incremental steps. Likewise, the MDA coverage in patch 2 is increased from 0% to 100% on the Y-axis, while the MDA coverage in patch 1 is fixed at a particular value. These permutations resulted in 10,100 simulations, summarized in the surface plots below.

These sets of 10,100 simulations are repeated for the MDA coverage values in patch 1 from 0% to 90% with 10% increments and for the identical, lower and higher pre-intervention disease intensities in patch 2 compared to patch 1.

## Results

For a particular malaria incidence, there exists a specific “baseline” or “minimal” threshold of MDA coverage above which elimination could be achieved in an isolated patch. In Figure 2, the “baseline” MDA threshold for patch 2 is at the horizontal red line which can be identified by the change from light-blue pixel to grey pixel, where the value on the connectedness (X-axis) is 0%. More generally, the required MDA coverage threshold for malaria elimination in patch 2 is given by the line separating the light-blue and dark-blue areas (where elimination is achieved) from the grey and orange ones, where elimination only happens in patch 1 or is not attained at all.

**Figure 2:**
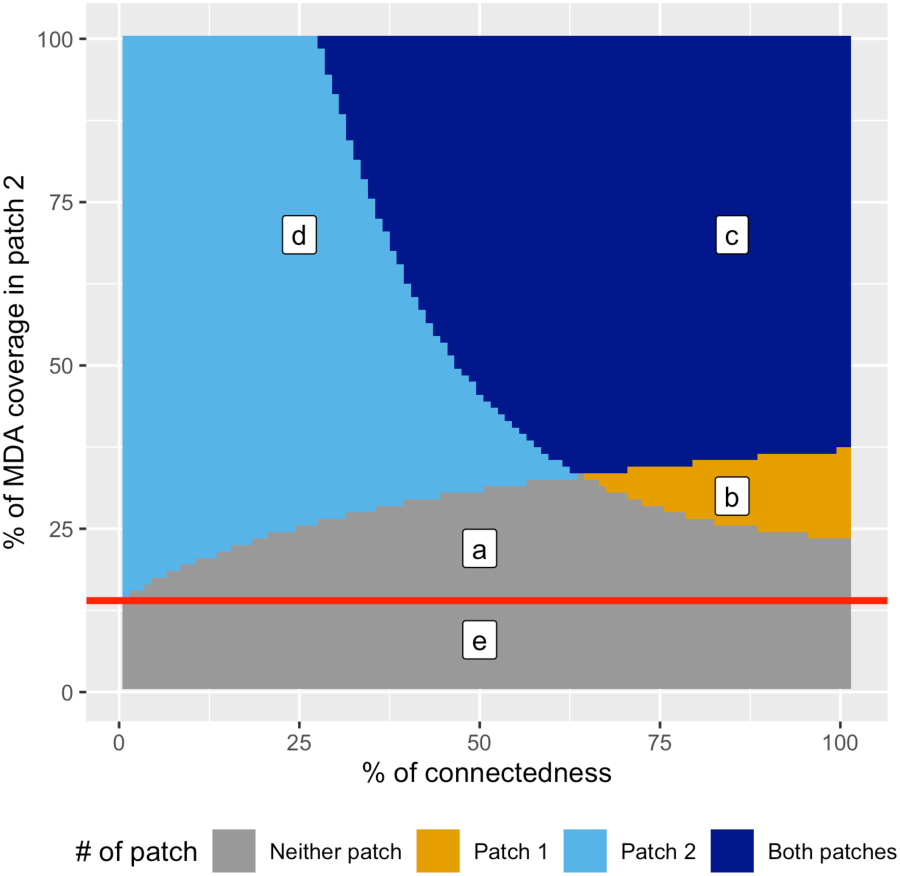
How to interpret the surface plots in the result. On the left edge of the plot where connectedness is 0%, take the point where the grey or the orange colour changes to the light blue or the dark blue colour. A horizontal line from that point (red line in the figure) is the “baseline” or “minimal” MDA threshold for the patch 2 to achieve elimination if it were isolated from other patches. Deviations from this line describe the assembly effect of patch 2. The joint areas a+b illustrate a negative assembly effect on patch 2, describing how an increase in MDA coverage would have to follow an increase in connectedness with another patch for elimination to be possible in patch 2. The joint areas b+c depict a positive assembly effect on patch 1, showing how an increase in connectedness with patch 2 can help eliminate transmission in patch 1 granted that it has a sufficiently high MDA coverage. This figure is merely an illustration and it is not an actual model result.

As connectedness increases, the MDA coverage threshold deviates from the “baseline”. This change in coverage threshold for a successful intervention in a patch depending on its connectedness to another patch is hereafter referred to as an “assembly effect”, which is closely related to a herd effect but accounts for connectedness between populations. This assembly effect can have either positive (i.e. protective) or negative implications for individuals in either patch. In Figure 2, a+b is the negative assembly effect for patch 2, where increasing connectedness with the patch 1 increases the MDA coverage threshold required for elimination in patch 2. From the point of view of the patch 1, b+c is the positive assembly effect – patch 1 doesn’t achieve elimination when it is isolated, but it does after a certain level of connectedness.

**Figure 3:**
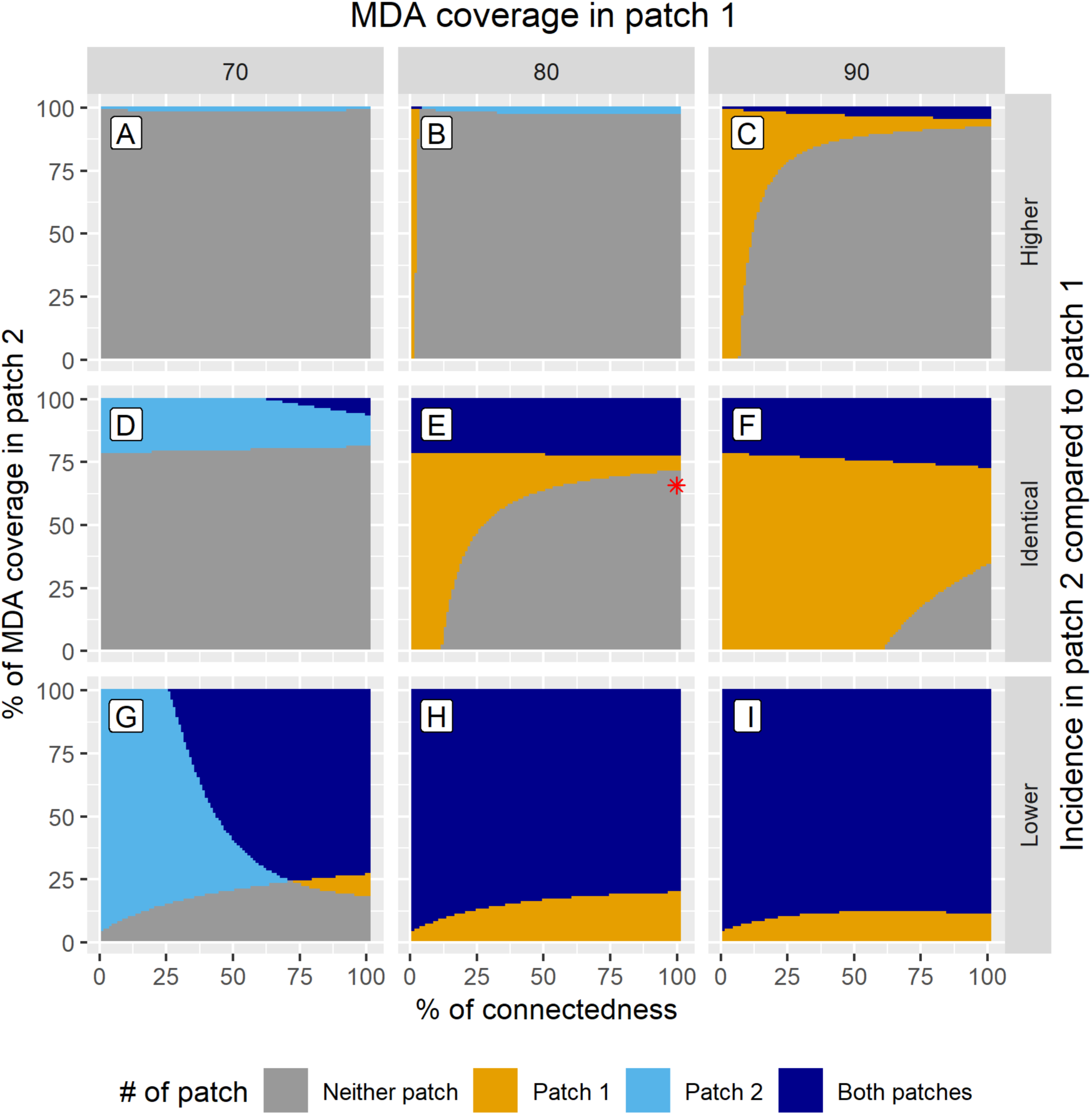
Achieving elimination in two connected patches by varying connectedness between the two populations (x-axis) and MDA coverage in the 2^nd^ patch (y-axis). Columns represent different sets of MDA coverage in patch 1 (70%, 80%, and 90%, respectively). Each row represents the relative incidence level between the two patches. The red askterisk represents the combination of parameter values matching the MDA trial implementation described in Parker et. al.

### Assembly effect between two patches with the same incidence

In the middle row of Figure 3, both patches have an identical pre-intervention incidence that implies a baseline MDA threshold of 78% coverage to achieve elimination if the villages were isolated. In Figure 3:D, there is a negative assembly effect for patch 2 (the grey area above the baseline MDA threshold) because of the increasing connectedness with patch 1 which has a low MDA coverage of 70%. However, the increasing connectedness is beneficial to patch 1 – elicited by a positive assembly effect. Despite patch 1 having 70% MDA coverage, and not being able to achieve elimination on its own, the increasing connectedness with patch 2 at 94% MDA coverage and above makes elimination attainable in patch 1 (dark blue triangle at the upper right corner).

A reciprocal effect is seen when patch 1 has higher MDA coverage that is enough to achieve elimination on its own (Figure 3: E and F). Patch 2 now experiences a positive assembly effect, inticated by the extension of the dark blue areas below the baseline MDA coverage threshold of 78%. Patch 1 experiences a negative assembly effect; as connectedness increased, elimination in patch 1 is not predicted to occur for low MDA coverage in patch 2 (grey area in the lower right corners).

**Figure 4:**
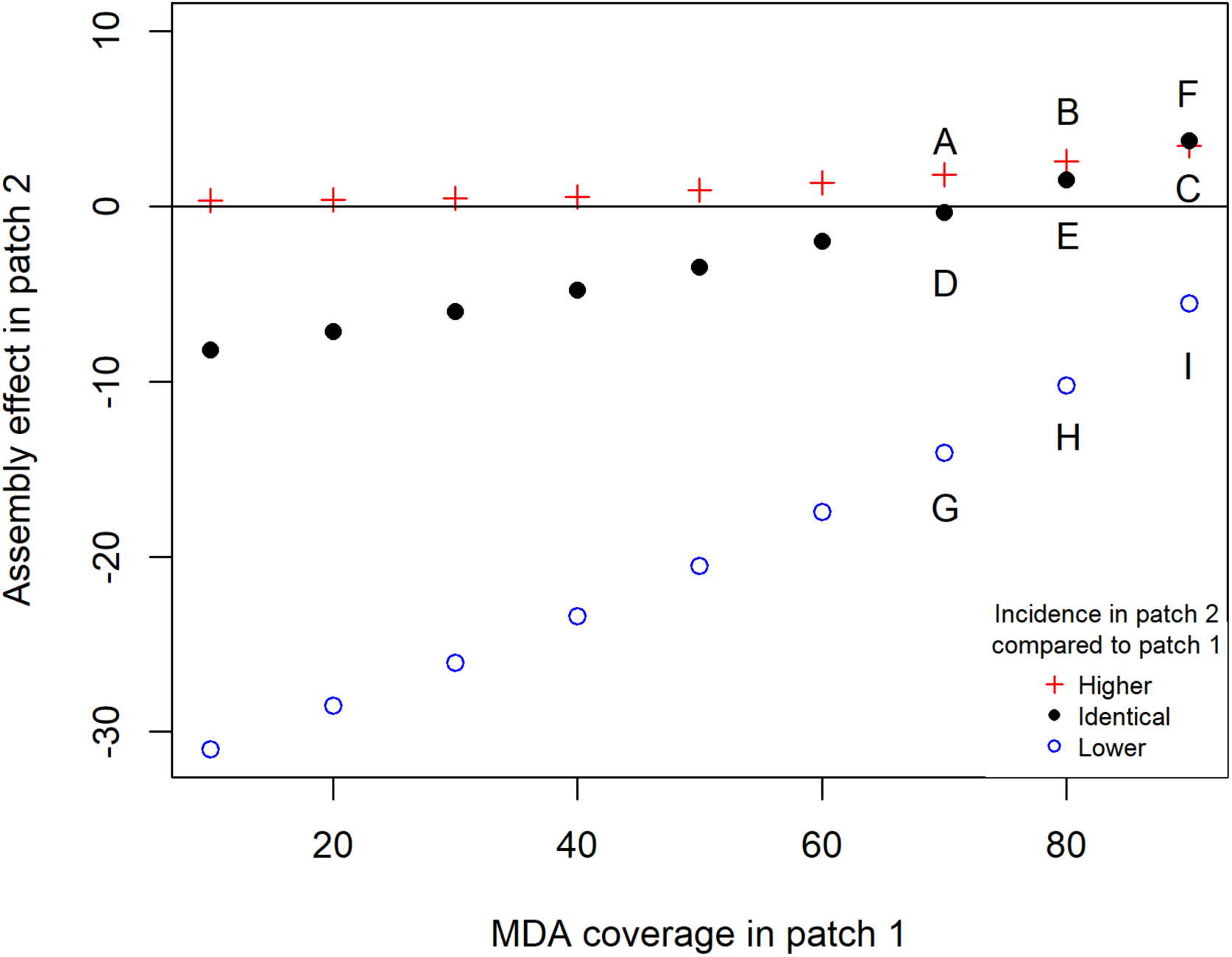
Assembly effects in patch 2 where relative incidence is higher, identical or lower compared to patch 1. The value of assembly effects on the X-axis was calculated by integrating the assembly effects in patch 2 over all level of connectedness with patch 1. A, B, C, D, E, F, G, H, and I represent the assembly effects of respective panels in Figure 3. Blue circles reporesent the assembly effects in non-hotspot for different coverage in the hotspot. Red crosses represent the assembly effects when incidence in patch 2 is so high that MDA is not an effective intervention (i.e. 100% MDA coverage is required to achieve elimination in patch 2). Black dots represent the assembly effects when the two patches have identical incidence (i.e. assembly effects are the same from the point of view of both patches).

Because the pre-intervention transmission intensities are the same in the two patches, the resulting assembly effects are purely due to differences in intervention coverage. The total assembly effect in patch 2 increases with increasing intervention coverage in patch 1 (black dots in Figure 4). The switch from negative assembly effect to positive assembly effect occurs at the “baseline” coverage threshold for the particular disease intensity shared by both of the patches.

We also compared the model’s prediction against results from an MDA trial described in Parker et al.^4^ where a village failed to achieve elimination presumably due to a cluster of non-participation in the MDA. This scenario was modelled as a set of two contiguous patches with 100% connectedness and with identical incidence. One patch received around 80% MDA coverage and the other, 64%, with the latter representing the non-participation cluster (details in supplementary materials). The model predicted that neither patch would achieve elimination (the red asterisk in Figure 3:E).

### Assembly effect when two patches have different incidences

#### Hotspot vs non-hotspot

In the bottom row of Figure 3, patch 2 has a 25% lower pre-intervention incidence compared to patch 1. This mimics a scenario where a low incidence community (nonhotspot: patch 2) is connected to a high incidence community (hotspot: patch 1). The MDA coverage threshold for elimination is very low at 5% for the non-hotspot, whereas it is 78% for the hotspot, when connectedness is zero.

When MDA coverage in the hotspot is slightly below its required threshold (Figure 3:G), we uncover both a negative assembly effect for the non-hotspot and a positive assembly effect for the hotspot simultaneously. This suggests that, when connectedness is over 60%, even if an optimal MDA coverage (i.e. 78%) is not achieved in the hotspot, elimination could be attained in both patches by increasing MDA coverage in the non-hotspot. For a 60% connectetdness, MDA coverage over 30% in the non-hotspot is predicted to result in elimination in both patches.

In panels H and I of Figure 3, the hotspot has an adequate MDA coverage at 80% and 90% respectively, and is predicted to always achieve elimination, regardless of the level of connectedness and the value of MDA coverage in the non-hotspot.

Figure 4 displays how the assembly effect in a particular patch is modulated by its connectedness to the other patch for different relative incidence ratios. Non-hotspots will always experience a negative assembly effect, albeit with a declining intensity for increasing coverage in the connected hotspot. The opposite is true for the hotspots (supplementary material: Figure 4). These trends suggest that the difference in transmission intensity is the main determinant of what types (positive or negative) of assembly effects can be observed.

In Figure 3:I, the required intervention threshold plateaus between 40% and 80% of connectedness. Further increase in the connectedness decreases the required intervention threshold slightly.

### Assembly effect when intervention is ineffective for the connected patch

An intervention may not be appropriate if the disease intensity is too high (e.g., MDA doesn’t work in a high-transmission setting). We simulated this scenario in the first row of Figure 3 by setting patch 2 as a high-transmission setting. In isolation, patch 2 would require a 100% coverage, and patch 1 would require more than 78% coverage of MDA for elimination to be attainable. As a consequence of being connected to patch 2, the prospects for elimination in patch 1 would be greatly diminished (large negative assembly effect in Figure 3: B and C).

## Discussion

In a single patch system, the success of an intervention depends on the pre-intervention disease intensity and the coverage of the intervention, provided the intervention is efficacious. In our two-patch connected system, whilst those metrics are still relevant, the level of connectedness between the two patches is a key determinant of the intervention’s success. Our results illustrate how connectedness can bring an advantageous effect to one patch, while potentially disadvantageous to the other. We have designated this effect the *assembly effect* and define it as:

*“The difference in the minimum intervention coverage required for a successful intervention in a specific patch when it is isolated versus when it is connected in some degree to another patch with potentially different disease intensity and/or different intervention coverage*.”

An assembly effect can be seen when connectedness is as low as 1%. Its magnitude and direction of effect depend on transmission intensity and intervention coverage in the adjacent area.

When connected patches have identical pre-intervention disease intensity, but different intervention coverage, the required threshold for a successful intervention in each patch will equilibrate with increasing connectedness. A negative assembly effect occurs in a patch when it is connected to another patch that doesn’t have enough intervention coverage to control its transmission intensity. A positive assembly effect occurs otherwise. Therefore, if one patch achieves a higher-than-optimal coverage of intervention, and its connected patch has a less-than-optimal coverage, it is still possible to attain a successful outcome in both patches. This has implications for public health interventions in locations with low adherence. In settings where multiple communities or populations are highly connected, as long as a certain number of the populations achieve higher-than-optimal coverage, the remaining populations can have less-than-optimal coverage.

As countries move towards elimination and transmission intensity distributions over space become extremely patchy,^8^ it becomes increasingly important to target hotspots with an adequate intervention coverage. Our results suggest that in order to achieve elimination, adjacent non-hotspot areas should not be left without interventions. Having some intervention coverage in the non-hotspots is also helpful when the optimal intervention coverage could not be achieved in the hotspots.^20^ We predict a significant positive assembly effect in hotspots with sub-optimal intervention coverage even when intervention coverages are modestly improved in the neighbouring non-hotspots (Figure 3:G and supplementary material: Figure 4).

Public health interventions that reduce transmission and target populations which are not in complete isolation will likely also result in an assembly effect. By taking consideration of connectedness between populations, overall disease intensity, and adherence to the public health intervention(s) being used, communicable diseases can more effectively be controlled and eliminated.

### Implications for the focal malaria interventions

The WHO has recommended MDA as a potential tool to accelerate malaria elimination and recommended its deployment only when core malaria interventions are already delivered in high quality coverage and the area where it is implemented is in very low transmission setting.^8^ Our result aligns with this recommendation by showing how it could be ineffective when applied before very low transmission is achieved in all connected patches. Once very low transmission is achieved in all connected patches through improvement and maintenance of core malaria interventions, some patches with relatively higher incidence (hotspots) and relatively lower incidence (non-hotspots) could persist. In such a scenario, it would be tempting to target the malaria hotspots with MDA. Our results suggest that targeting only the malaria hotspots may not be enough. It is often challenging and resource intensive to achieve high coverage for MDA,^21,22^ and the imported asymptomatic infections from the connected non-hotspots could refuel transmission.^23^ Therefore, when targeting hotspots in these scenarios, reinforcement of interventions in adjacent non-hotspots would benefit the hotspots because of the positive assembly effect and improve the chance of a successful elimination campaign. An example guideline for malaria elimination is described in Table 1.

**Table 1.** Example guidelines for a malaria elimination scenario.

|  |  |
| --- | --- |
| Background scenario | Suppose we're planning to eliminate malaria from a province with very low malaria transmission. |
| Adequate core malaria interventions and identification of malaria hotspots | First, we have to ensure the quality coverage of core malaria interventions such as early diagnosis and treatment, and long-lasting insecticide-treated nets (LLINs) in all villages within the province. We then need to identify the hotspot villages based on prevalence surveys or incidence reports. |
| Information on connectedness | Depending on the budget and the available timeframe, connectedness between villages can be inferred in several ways. Remote sensing and GIS analysis may be used to infer connectedness through metrics such as distance, estimated population size, and estimated travel time. Human mobility surveys maybe conducted to inform connectedness. GPS logger studies maybe more expensive and labour intensive, but could produce more detailed measures of connectedness. |
|  | A multi-patch or individual-based model maybe used to fit historical data of a similar area to yield an estimate of the connectedness. |
| Optimisation of intervention coverage across hotspots and non-hotspots | Armed with some information on the connectedness between villages and the location of hotspots, we can strategize to ensure efficiency and effectiveness of the focal MDA is optimised. All malaria hotspots should aim to reach an MDA coverage over the minimal threshold (i.e. 80% in most context). Non-hotspot villages that are connected to the hotspots should get an MDA coverage of at least 30%. |
| Example calculation of MDA rounds required for the intended effective coverage | The MDA coverage which we have used here is the percentage of the target population who receives at least one round of MDA. Different total coverage levels could represent different number of monthly rounds of MDA. In our model, the final MDA coverage of x% after 3 rounds means $1-(1-x)^{(1/3)}$ coverage in the round 1. Therefore, if we achieve 70% of total MDA coverage after 3 rounds, we could say that 1 round of MDA will cover 33% of the total population. MDA coverage from our model can thus be operationalized into number of MDA rounds. Using this information in our example scenario would mean that we could target malaria hotspots with |
|  | three rounds of MDA while the non-hotspots which are connected to the hotspots could be provided with only one round of MDA. |

### Limitations

This model was developed as a theoretical framework to define the concept of the assembly effect in a general sense. There were many assumptions in the model structure and parameter values used. The way MDA was modelled in the compartmental system may not be an accurate representation of a real-world MDA. The model has so far been validated on a single scenario. Further rigorous validation and fitting would be required to use it as a predictive tool. The time point for measuring the outcome was arbitrarily set as one year after the completion of MDA. Results will vary depending on where this time point is set.

## Conclusions

Assembly effect is a population version of the herd effect and it occurs between connected populations of potentially different disease intensities and/or intervention coverages. The ultimate impact of an intervention in an area depends on how well it is connected with neighbouring areas. Information on the level of connectedness between populations will inform efficient control and elimination strategies. For malaria, improving and maintaining core malaria interventions is the first step towards achieving very low transmission, which could be followed by an acceleration to elimination. In implementing the accelerating activities such as MDA, targeting malaria hotspots alone may not be optimal. Having the positive assembly effects on the hotspots by additionally implementing MDA with lower coverage on their connected non-hotspots will lower the required MDA coverage threshold in the hotspots and will increase the feasibility of malaria elimination.

## Data Availability

The curated programming code for all the experiments are freely accessible on Github at this address: https://github.com/SaiTheinThanTun/MDA_eff_pub

## Authors’ contributions

STTT: Conceptualization, Model development and simulations, Drafting and revising the manuscript

DMP: Conceptualization, Revising the manuscript

RA: Model development, Revising the manuscript

LJW: Conceptualization, Model development, Revising the manuscript

